# Cost-effectiveness of a UK-based primary healthcare intervention: Improving Medicines use in People with Polypharmacy in Primary Care (IMPPP)

**DOI:** 10.1101/2025.10.23.25338657

**Authors:** Nouf S. Gadah-Jeynes, Ammar Annaw, Rupert A Payne, Deborah McCahon, Jeff Round

**Affiliations:** Department of Population Health Sciences, Bristol Medical School, University of Bristol, Bristol, UK; Exeter Collaboration for Academic Primary Care, University of Exeter, Exeter, UK; Institute of Health Economics, Edmonton, AB, Canada; Faculty of Medicine and Dentistry, University of Alberta, Edmonton, AB, Canada

**Keywords:** Polypharmacy, medication review, cost-effectiveness, primary care

## Abstract

The prescribing of multiple medicines to one individual, or polypharmacy, is increasingly common. While the use of multiple medications by a patient is often appropriate, in some cases prescribed medicines may not have the intended benefit and may even cause harm, and it is important to understand the clinical and economic implications of polypharmacy and interventions to optimise prescribing. In the UK, most ongoing clinical management of polypharmacy takes place in primary care. We estimated the cost-effectiveness of the Improving Medicines use in People with Polypharmacy in Primary Care (IMPPP) trial from a UK NHS perspective. IMPPP was a pragmatic, open-label, two-arm cluster-randomised trial across 37 English general practices, including 1,715 patients. The intervention comprised a structured, enhanced process for delivering patient-centred polypharmacy reviews, and was compared to control arm practices delivering usual care. Costs were derived from routine electronic health records including primary and secondary care service utilisation data whilst QALYs were estimated via SF-12v2. Follow-up was assessed at 6 months compared to pre-randomisation baseline. Cost-effectiveness was assessed using multilevel modelling with bias-corrected and accelerated bootstrapping to calculate 95% confidence intervals. Additional one-way sensitivity analyses were conducted to explore uncertainty. Adjusted mean QALYs were slightly higher in the intervention group (0.629) versus control (0.624), with a non-significant difference of 0.006 (95% CI: -0.002 to 0.014). Mean adjusted costs were also higher in the intervention group (£4166 vs. £3655), with a non-significant cost difference of £511 (95% CI: -£73 to £949). The probability of cost-effectiveness at National Institute for the Health and Care Excellence’s £20,000/QALY and £30,000/QALY thresholds were 6% and 12% respectively. Complete case analysis showed a £138 NHS cost reduction (95% CI: -£652 to £376) and a QALY gain of 0.012 (95% CI: 0.004 to 0.021). Polypharmacy medication review as conducted in the IMPPP trial is not cost-effective. This probably reflects multiple factors, including clinical effectiveness outcomes and key cost outcomes being relatively insensitive to the intervention.

## 2. Introduction

Polypharmacy is broadly defined as the prescribing of multiple medicines to one individual (1). Although there is no consensus on how many medicines constitute ‘multiple’, polypharmacy is increasingly common irrespective of how it is measured due to an ageing population and increasing multimorbidity (1, 2). Given that most prescribing occurs in primary care, where people with long-term conditions are increasingly managed, polypharmacy presents a particular challenge to general practice (GP) (1). Although polypharmacy has typically been considered as something to avoid (3), there is a need to distinguish between appropriate and problematic polypharmacy (3). Appropriate polypharmacy occurs where medication use is optimised, while problematic polypharmacy occurs where medications are used inappropriately or where the intended benefit is not realised (3). Medication optimisation strategies for managing polypharmacy therefore need to balance expected benefits and risks with patient goals and priorities and should ideally not only demonstrate reductions in potentially inappropriate prescribing but also benefits to patient outcomes (1). Recent national guidance on optimising care for polypharmacy has been produced in the UK, but it is not supported by high quality evidence of intervention effectiveness and cost-effectiveness (4–6). Furthermore, there has been significant recent investment in pharmacists working in UK general practice to improve effective and safe use of medicines (7), but again evidence is lacking on how best this resource can be utilised in the context of polypharmacy (1).

## 3. Methods

### 3.1 Overview of economic evaluation

We conducted cost-utility and cost-effectiveness analyses alongside the IMPPP trial (1). The trial investigated the clinical and cost-effectiveness of a targeted medication review for patients with polypharmacy in primary care. A health economic analysis plan (HEAP) was developed and published prior to analysis (1, 8). The primary analysis took an NHS and Personal Social Services (PSS) perspective, while a secondary analysis used a societal perspective, as per the National Institute for the Health and Care Excellence’s (NICE) guidelines (9). All research regulatory approvals were obtained, and all patients provided their consent to participate (1). Reporting of this analysis adheres to the Consolidated Health Economic Evaluation Reporting Standards (CHEERS) (see Supplementary materials, Table S1).

### 3.2 Participants

Eligible participants were adults aged ≥18 years with capacity to consent and registered with a participating practice. Recruitment targeted up to 50 patients per practice. Polypharmacy was defined as taking at least five regular medicines listed on “repeat” prescriptions, regardless of the last issuing date and flagging at least one indicator of potentially inappropriate prescribing (PIP) on automated case-finding (1). Polypharmacy in this trial was not specific to a condition or a disease area but instead was a generic medication review that included 118 PIP indicators over 15 clinical areas (1); our patient group therefore had a wide range of clinical profiles. This included variation in acute and chronic care needs, as well as age groups, reflecting real-world settings. PIP refers to “the prescription of potentially inappropriate medications associated with a higher risk of adverse outcomes such as drug interactions, falls, and cognitive impairment” (10, 11). A typical PIP example used in the trial was “Aged 65 or older and on drugs that cause constipation and on a laxative”; the full list of PIP indicators is published in the protocol (1). “Repeat” medicines are those where patients are able to request a refill from their practices without further consultation with a clinician (1).

### 3.3 Trial design and setting

IMPPP was a multicentre, open-label, cluster-randomised controlled trial with two parallel groups, aimed at assessing the effectiveness and cost-effectiveness of a targeted medication review versus usual care (control). The trial was carried out across 37 general practices in England. Detailed trial methods are available elsewhere (1). Eligible participants were identified using an informatics tool that searched GP electronic records to identify participants with multiple medications (polypharmacy) and PIPs (1). The tool was applied to participating practices’ patient lists after practice staff received training on the trial protocol, but prior to practice randomisation (timepoint T_1_). Practices were randomised in pairs (one usual care, one intervention) with regional stratification (Bristol, West Midlands) (1). Key time points were as follows: 6-month period prior to practice randomisation (timepoint T_0_); practice randomisation (T_1_); pre-review eligibility confirmation for all participants (T_2_) which was immediately followed by a medication review (intervention arm only); and follow-up (T_3_), six-months after T_2_. Medication review delivery to the 50 participants per practice occurred over a 6-month period. Variable logistical delays in the delivery of training post-T_1_ and the fact that reviews were delivered spread over a 6-month period, led to expected differences in time between T_1_ and T_2_ amongst practices and patients (1).

### 3.4 Intervention and comparator

The intervention is a service model based in primary care. A comprehensive description of the intervention is reported in the protocol (1). Briefly, the intervention was based on a five-stage review process automated case-finding (stage 1), notes review by a pharmacist (stage 2), pharmacist-GP liaison (stage 3), a patient-centred structured clinical approach to medication review (stage 4) and a follow-up appointment as deemed clinically appropriate (stage 5) (1). The intervention also included components to enhance professional engagement (clinician training, practice feedback and financial incentives), Figure 1 (1). An informatics tool integrated into GP clinical systems helped support structured case-finding, the medication review itself, and the practice feedback component (1). Usual care comprised access to routine medication reviews as part of standard clinical practice, but no specific management strategy focused on polypharmacy (1).

**Figure 1:**
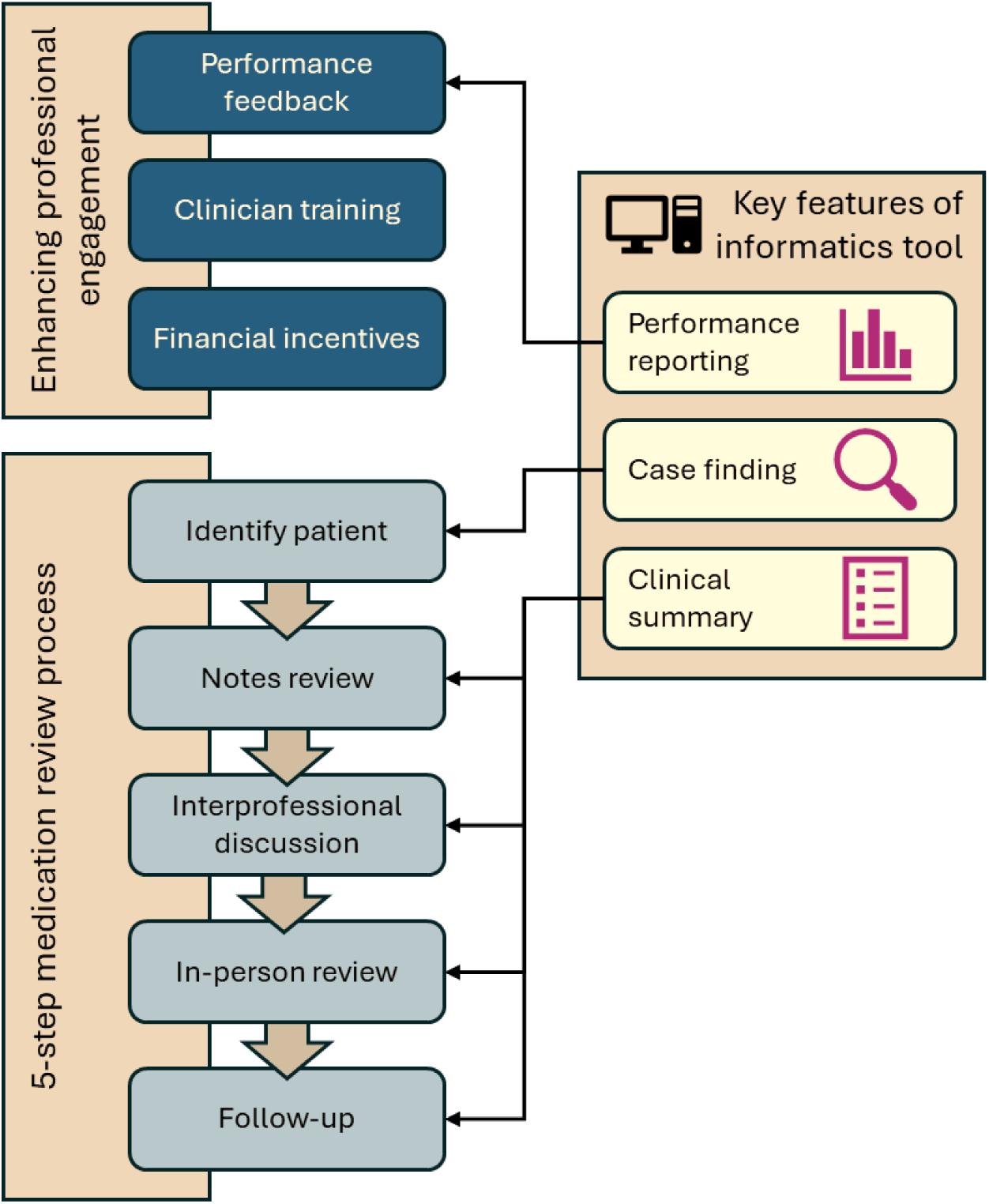
intervention components.

### 3.5 Time horizon

The economic evaluation was conducted alongside the IMPPP trial. The primary clinical effectiveness outcome was the number of PIP indicators at six months, which was deemed an appropriate time-horizon to detect a clinical improvement. The time horizon of the economic evaluation was chosen to mirror the primary study timeline and the study was powered accordingly. In usual care, medications review generally occur at least annually, but may happen more frequently for high-risk groups (e.g. patients in care homes, those with more complex and problematic polypharmacy) or as part of routine care of patients with specific long-term conditions (e.g. diabetes, asthma) (12). However, the IMPPP intervention was designed around delivery of a single review, and extending the time horizon to 12 months or beyond could have resulted in capturing more than one review and been challenging to interpret.

### 3.6 Outcome measurement

The outcome for the primary economic analysis was Quality Adjusted Life Years (QALYs). Participants were asked to complete the Short-Form 12-Item Survey-version 2 (SF-12v2) and a 0–100-point visual analogue scale (adapted from the EuroQol Visual Analogue Scale, EQ-VAS) at T_1_, T_2_ and T_3_. Participants’ SF-12 responses were mapped to utility scores using PROCoRE v2.4 (QualityMetric, Johnston, RI, USA). The area under the curve method was used to convert utility scores into QALYs (12). For participants who died during the follow-up, we assigned a utility value of zero from the date of death onwards. For the secondary analysis, we report the adjusted incremental cost-effectiveness of a PIP indicator avoided. As part of an external pilot phase, an investigation of the appropriateness of the EQ-5D-5L and SF12v2 was conducted. As part of the pilot work, respondents completed both instruments so that we could evaluate suitability for inclusion in the full study (the IMPPP trial). One key indicator we measured was response rate and data-completeness for each instrument, and we found no statistically significant differences between them during the pilot. Given prior evidence that the SF-12-v2 could possibly be more sensitive to health status changes as a result of polypharmacy and changes in prescribing, we chose the SF-12-v2 for use in the IMPPP trial (14).

### 3.7 Resource-use data collection, coding, and valuation

The primary economic analysis was conducted from the National Health Service and Personal Social Services(NHS and PSS) perspective as suggested by NICE (9) Primary Electronic Health Record (EHR) and secondary care administrative data were obtained for the period T_0_ to T_3_. Intervention-related appointments between clinicians and participants, or interprofessional meetings between a GP and a pharmacist, were captured in the EHR. Primary care EHRs included all primary care contacts with GPs, nurses, Allied Health Professionals (AHPs), pharmacists and administrative staff that were conducted either in-person at the practice, virtually (telephone/video calls/emails), or as home visits. Primary care EHRs were also used to obtain data on prescribed medications. Medications were costed using the prescription cost analysis (PCA) (2022/23). The PCA provides details of the number of items and the Net Ingredient Cost (NIC) of all prescriptions dispensed in the community in England (15). We used a weighted average by medication using the average prescription cost from the cost per quantity column in the Presentations tab of the published spreadsheet.

The intervention was anticipated to include essential ‘non-patient-facing’ tasks associated with case finding and clinical discussions between GPs and pharmacists. As such tasks were conducted by both administrative and clinical staff, they were incorporated into the analysis and categorised as a “non-patient-facing” activity. Primary care data was provided from practices using two clinical Information Technology (IT) systems: EMIS (n=35) and SystmOne (n=2). Secondary care data was based on Hospital Episode Statistics (HES), and included Accident and Emergency (A&E) visits, inpatient admissions (elective day cases, non-elective short and long stays) and outpatient visits. Social services provided by local governments included Meals on Wheels (MoW), attending day care centres, home care help services and social worker visits; these were captured through self-report questionnaires at T_1_, T_2_ and T_3_ and were reported under the public sector perspective (i.e. NHS cost + local government costs for social services) In our societal perspective analysis, we included out of pocket health care cost (travel cost, over-the-counter medications cost and private treatments) and under this perspective, we reported public sector costs and out of pocket payments by patients. The NHS cost inflation index was used to inflate unit costs to current prices as necessary (13). Table S2 outlines how the resources were measured, coded and valued using 2021–2022 UK £ prices. For participants who died during the follow-up period (T_2_ to T_3_) costs were assumed to be zero from the date of death onwards. For each participant, total NHS costs were calculated as the sum of all primary and secondary care costs, prescribed medications cost and the intervention cost (Table 1 lists how the intervention was costed).

**Table 1:** Intervention-related costs.

|  | **Cost** | **Note** |
| --- | --- | --- |
| ***Informatics tool*** |  |  |
| Software licence | £168.65 per practice in both arms. | Includes licence fees and helpdesk support. A per patient cost was calculated by dividing £168.65 by the practice size. |
| Additional server capacity | £985 per practice in both arms. | 17 out of 37 practices (45.95%) required additional server capacity. A per patient cost was calculated by dividing £985 by the practice size. |
| Performance feedback reports | £324 over 6 months. | This was completed by the trial team but is expected to be either performed at a practice-level, a PCN-level or at a national level. We estimated the cost for a practice manager (45 minutes a month). Estimated as total cost of generating reports/number of patients recruited in the intervention arm only. |
| ***Clinical training*** |  |  |
| Initial clinician training | £264 for 4 hours | 4 hours per pharmacist per practice. A cost per patient was calculated by dividing the total cost of training by the number of patients recruited in the intervention arm only. |
| Refresher training | GP:<br>£479 (for two hours)<br>Pharmacist:<br>£132 for 2 hours | 2 hours per GP per practice.<br>2 hours per clinical pharmacist per practice.<br>A cost per patient was calculated by dividing the total cost of training by the number of patients recruited in the intervention arm only. |
| ***Financial incentives*** | £60 per patient | For delivering notes review, interprofessional discussion, and in-person review and a follow-up unless exempted (Figure 1). |
| ***Reimbursement*** |  | Captured in electronic health records. |

### 3.8 Analysis

Stata/ IC version 19.0 or higher (StataCorp) was used for all analyses. The primary economic analysis was to evaluate cost and outcomes from T_1_ to T_3_ whilst accounting for baseline outcomes (T_0_). The periods between ‘T_0_ to T_1_’ and between ‘T_2_ to T_3_’ are fixed 6-month periods, unlike the period from T_1_ to T_2_ where there were variations between practices in completing training and reviewing participants; therefore, the baseline for the analysis was taken as the 6-months prior to T_1_.

The analysis was based on the intention-to-treat (ITT) approach (14). Analyses were adjusted for baseline characteristics determined at T_1_, including age (continuous), sex (binary), Index of Multiple Deprivation (IMD) decile (1st decile is most deprived, 10th decile is least deprived), Cambridge Multimorbidity Score (CMS) (continuous), geographical region (Bristol or West Midlands), and number of distinct medicines issued within 84 days prior to T_1_ (continuous). Other variables controlled for in the analysis include time between T_1_ and T_3_ (continuous), intervention allocation (binary), and baseline cost (T_0_ to T_1)_ and utility (T_1_) values as appropriate. A mixed-effects multilevel model (MLM) was used with GP practices fitted as a random effect and all other covariates fitted as fixed-effects. It has been found that MLM performs better than ordinary least squares (OLS) regression in trial-based economic evaluation where the data was clustered (15). The MLM was used to calculate the adjusted mean costs and QALYs and the adjusted mean differences in costs and QALYs between groups. The 95% confidence intervals (CIs) were estimated using bias-corrected and accelerated bootstrapping. The primary economic analysis estimated the total NHS/PSS costs and QALYs of the intervention and control groups, and the between group difference in costs and QALYs. Results are reported as incremental cost-effectiveness ratios (ICER) and as an incremental net-monetary benefit (iNMB). The iNMB is presented from the NHS perspective assuming a willingness to pay (WTP) threshold of £20,000 per QALY gained (9). Discounting was not applicable due to the time horizon being less than a year-. In the cost-effectiveness analysis, we estimated the cost per PIP indicator avoided at T_3_ and adjusted for the same variables as in the primary analysis.

One-way sensitivity analyses were conducted to explore key methodological uncertainties associated with missing data (described below) whilst adjusting for the same covariates as per the main analysis:

- Available cases only which included participants with complete costs and QALYs.
- Additional adjustment for IT informatics systems: EMIS or SystmOne (binary).
- Additional adjustment for mental health problems: Anxiety/depression (binary), alcoholism (binary) and severe mental illness (binary) that were captured within the Cambridge Multimorbidity Score (16)
- Analysing outcomes between T_2_ and T_3_ (QALYs and costs) whilst adjusting for T_2_ outcomes instead of T_1_ outcomes (in case intervention effect might vary over time, post-randomisation).

We used the output from the MLM models to estimate the incremental net monetary benefits (iNMB) statistics and associated 95% CIs at NICE’s recommended WTP threshold of £20,000/QALY (9). We constructed a cost-effectiveness acceptability curve (CEAC) to present the uncertainty over a range of WTP threshold. We also used the output from the MLM model to estimate the cost per PIP indictor avoided.

### 3.9 Missing data and sensitivity analyses

A small proportion of primary care contacts (2.67%, n=3201 observations) were missing details on the type of contact and were coded as ‘administrative activity’ in the primary analysis in agreement with the study’s principal investigator (RP).

All remaining missing data was assumed to be missing at random (MAR). MAR values were estimated using multiple imputation (MI) by chained equations with predictive mean matching. In the MI model (NHS and PSS perspective analysis model), we included contact and cost variables by contact type (e.g. in-person, virtual) for each HCP. In addition, intervention cost, utility/VAS scores, and secondary care resource use were included in the model as well as participants’ and practices’ characteristics. A separate MI model was carried out for the societal perspective analysis and included the same variables specified in the primary NHS perspective analysis in addition to costs of MoW, attending day care centres, home care help services and social worker visits. A series of sensitivity analyses were conducted to explore the impact of the assumptions used to address missing data.

## 4. Results

### 4.1 Descriptives

Data was available for 1715 participants recruited from 37 GPs in the Southwest (n=18) and West Midlands (n=19) in England. A total of 827 participants were recruited to the control group (West Midlands=442, Southwest=385) and 888 to the intervention group (West Midlands=560, Southwest=328). There were 25 participants who died during the trial (T_2_ to T_3_). Participants’ baseline demographics by group are provided in Table 2. The mean duration spent in the trial was 544 days (SD=59) for the intervention group and 536 days (SD=63) in the control group. On average, the intervention group experienced a longer delay between practice randomization (T_1_) and pre-review eligibility confirmation (T_2_), with a mean difference of eight days (95% CI = -13.96 to -2.38, p=0.0057) (Table 2). The number of PIP indicators at each time point was slightly higher in the intervention group than in the control group (Table 2). More people reported living alone in the intervention group (28%) compared with the control group (24%). Baseline utility scores (T_1_) did not significantly differ between the intervention and the control groups (Table 3), nor did their baseline unadjusted NHS cost (Table 4)., Men had significantly higher baseline utility scores than women (0.679 vs. 0.621), with a mean difference of 0.058 (95% CI= 0.043 to 0.072), but there was no significant difference in their baseline unadjusted NHS costs (men= £1916.08 ± £3322.73 ; women £1914.80 ± £2985.68), with a mean difference of £1.28 (95% CI= £-298.36 to £300.92).

**Table 2:** Participants’ baseline characteristics.

| Characteristics | Intervention (n=888) | Control (n=827) |
| --- | --- | --- |
| Number of practices | 19 | 18 |
| Age, mean±SD, years | 71.52±10.58 | 71.86±11.07 |
| Male, n (%) | 461 (51.91) | 416 (50.30) |
| Ethnicity – White, n (%) | 834 (96.53) | 781 (97.99) |
| Overall Cambridge comorbidity score, median (IQR) | 2.35 (1.42) | 2.38 (1.46) |
| IMD, n (%) |  |  |
| 1- most deprived | 30 (3.38) | 33 (3.99) |
| 2- | 69(7.77) | 46 (5356) |
| 3- | 61 (6.87) | 66 (7.98) |
| 4- | 101 (11.37) | 65 (7.86) |
| 5- | 96 (10.81) | 53 (6.41) |
| 6- | 65 (7.32) | 64 (7.74) |
| 7- | 134 (15.09) | 87 (10.52) |
| 8- | 116 (13.06) | 140 (16.93) |
| 9- | 94 (10.59) | 102 (12.33) |
| 10- least deprived | 122 (13.74) | 171 (20.68) |
| Living situation – alone, n (%) | 237 (28.05) | 194 (24.40) |
| Employment – fully retired, n (%) | 440 (76.39) | 403 (77.06) |
| Qualifications, n (%) |  |  |
| No education | 206 (23.20) | 195 (23.58) |
| GCSE/A Level/Diploma | 380 (42.79) | 355 (42.93) |
| University or higher | 161 (18.13) | 149 (18.02) |
| Median (IQR) duration between T<sub>1</sub> and T<sub>2</sub> in days<sup>a</sup> | 177 (74) | 160 (100) |
| Mean±SD duration of the trial in days<sup>b</sup> | 544.11±59.06 | 535.94±63.19 |
| Mean±SD number of PIP indicators per patient |  |  |
| T<sub>1</sub> | 2.39±1.83 | 2.30±1.73 |
| T<sub>2</sub> | 2.39±1.93 | 2.28±1.84 |
| T<sub>3</sub> | 2.30±1.93 | 2.29±1.82 |
| Total NHS cost, mean±SD | £5946.80±£5966.24 | £5627.16±£6486.05 |

<sup>a</sup> T<sub>1</sub> practice randomisation date; T<sub>2</sub>: pre-medications review and eligibility checks.
<sup>b</sup> Difference between T<sub>3</sub> (6-months post T<sub>2</sub>) and T<sub>0</sub> (6-months prior to T<sub>1</sub>).

**Table 3:** Mean unadjusted utility scores, VAS scores and QALYs for both groups based on available cases.

|  | **Intervention**<br>**(N) mean±SD** | **Control**<br>**(N) mean±SD** | **Mean difference<sup>a</sup>**<br>**(95% CI)** | **P value** |
| --- | --- | --- | --- | --- |
| *Utility scores* |  |  |  |  |
| T<sub>1</sub> | (828)<br>0.655±0.148 | (770)<br>0.645±0.149 | 0.010<br>(-0.004 to 0.025) | 0.173 |
| T<sub>2</sub> | (629)<br>0.665±0.146 | (565)<br>0.646±0.150 | 0.019<br>(0.002 to 0.036) | 0.027 |
| T<sub>3</sub> | (604)<br>0.642±0.174 | (557)<br>0.630±0.170 | 0.012<br>(-0.007 to 0.032) | 0.221 |
| *VAS scores* |  |  |  |  |
| T<sub>1</sub> | (861)<br>61.91±21.80 | (807)<br>60.13±22.44 | 1.79<br>(-0.340 to 3.911) | 0.100 |
| T<sub>2</sub> | (621)<br>61.73±22.00 | (562)<br>59.05±22.09 | 2.68<br>(0.163 to 5.200) | 0.037 |
| T<sub>3</sub> | (625)<br>60.30±22.10 | (573)<br>58.50±21.66 | 1.79<br>(-0.691 to 4.277) | 0.157 |
| QALYs<sup>b</sup>, mean±SD | (496)<br>0.653±0.170 | (428)<br>0.615±0.167 | 0.024<br>(0.008 to 0.040) | 0.003 |
| Imputed QALYs<sup>b</sup>, mean±SD | (496)<br>0.639±0.169 | (428)<br>0.613±0.169 | 0.020<br>(0.007 to 0.033) | 0.002 |

<sup>a</sup> unadjusted mean difference between the intervention and the control groups.
<sup>b</sup> adjusted for baseline utility scores only.

**Table 4:**
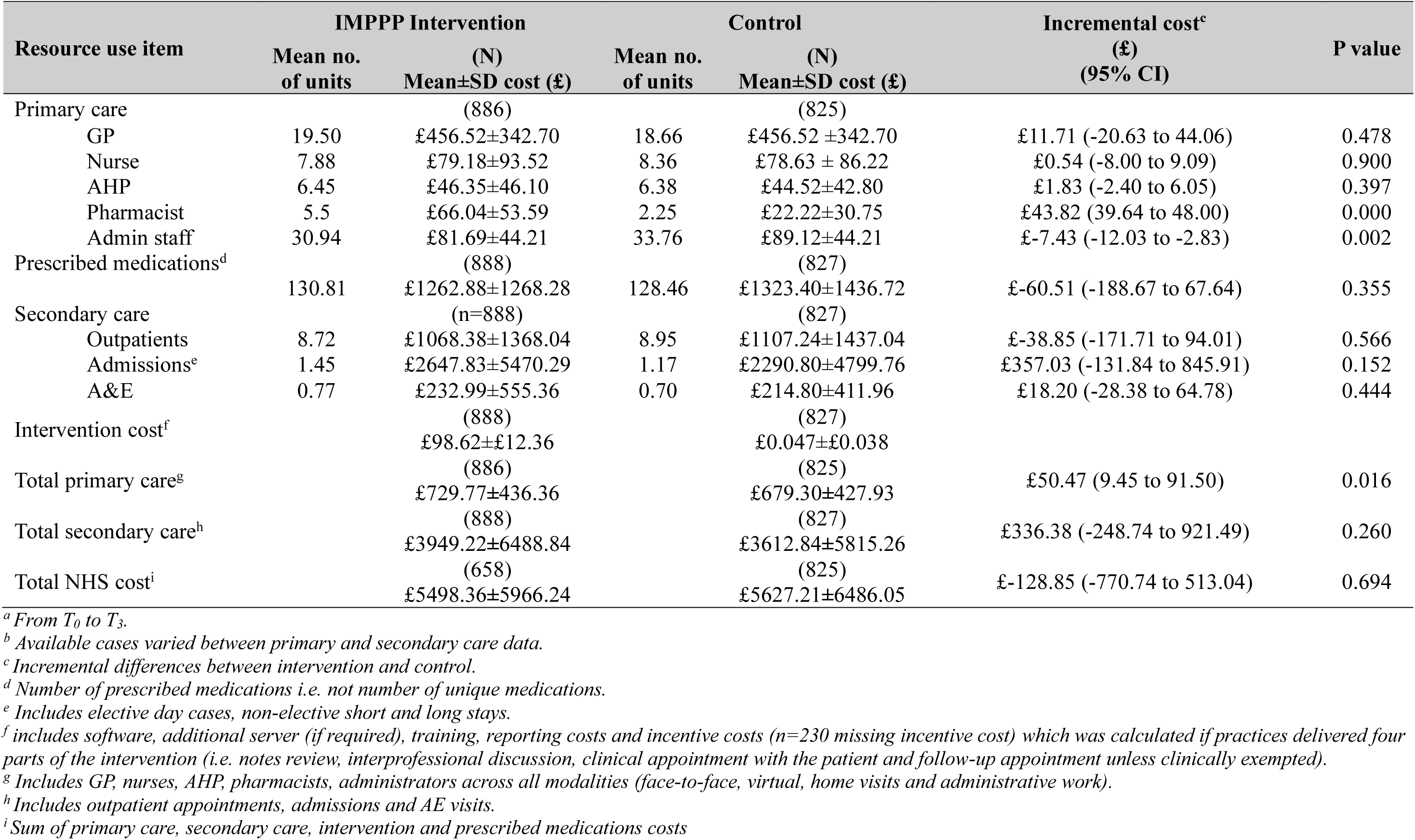
Mean unadjusted resource use contact and cost during the trial period^a^ in both groups based on available cases^b^.

| Resource use item | IMPPP Intervention |  | Control |  | Incremental cost<sup>c</sup><br>(£)<br>(95% CI) | P value |
| --- | --- | --- | --- | --- | --- | --- |
|  | Mean no.<br>of units | (N)<br>Mean±SD cost (£) | Mean no.<br>of units | (N)<br>Mean±SD cost (£) |  |  |
| Primary care |  | (886) |  | (825) |  |  |
| GP | 19.50 | £456.52±342.70 | 18.66 | £456.52 ±342.70 | £11.71 (-20.63 to 44.06) | 0.478 |
| Nurse | 7.88 | £79.18±93.52 | 8.36 | £78.63 ± 86.22 | £0.54 (-8.00 to 9.09) | 0.900 |
| AHP | 6.45 | £46.35±46.10 | 6.38 | £44.52±42.80 | £1.83 (-2.40 to 6.05) | 0.397 |
| Pharmacist | 5.5 | £66.04±53.59 | 2.25 | £22.22±30.75 | £43.82 (39.64 to 48.00) | 0.000 |
| Admin staff | 30.94 | £81.69±44.21 | 33.76 | £89.12±44.21 | £-7.43 (-12.03 to -2.83) | 0.002 |
| Prescribed medications<sup>d</sup> |  | (888) |  | (827) |  |  |
|  | 130.81 | £1262.88±1268.28 | 128.46 | £1323.40±1436.72 | £-60.51 (-188.67 to 67.64) | 0.355 |
| Secondary care |  | (n=888) |  | (827) |  |  |
| Outpatients | 8.72 | £1068.38±1368.04 | 8.95 | £1107.24±1437.04 | £-38.85 (-171.71 to 94.01) | 0.566 |
| Admissions<sup>e</sup> | 1.45 | £2647.83±5470.29 | 1.17 | £2290.80±4799.76 | £357.03 (-131.84 to 845.91) | 0.152 |
| A&E | 0.77 | £232.99±555.36 | 0.70 | £214.80±411.96 | £18.20 (-28.38 to 64.78) | 0.444 |
| Intervention cost<sup>f</sup> |  | (888) |  | (827) |  |  |
|  |  | £98.62±£12.36 |  | £0.047±£0.038 |  |  |
| Total primary care<sup>g</sup> |  | (886) |  | (825) |  |  |
|  |  | £729.77±436.36 |  | £679.30±427.93 | £50.47 (9.45 to 91.50) | 0.016 |
| Total secondary care<sup>h</sup> |  | (888) |  | (827) |  |  |
|  |  | £3949.22±6488.84 |  | £3612.84±5815.26 | £336.38 (-248.74 to 921.49) | 0.260 |
| Total NHS cost<sup>i</sup> |  | (658) |  | (825) |  |  |
|  |  | £5498.36±5966.24 |  | £5627.21±6486.05 | £-128.85 (-770.74 to 513.04) | 0.694 |

<sup>a</sup> From T<sub>0</sub> to T<sub>3</sub>.
<sup>b</sup> Available cases varied between primary and secondary care data.
<sup>c</sup> Incremental differences between intervention and control.
<sup>d</sup> Number of prescribed medications i.e. not number of unique medications.
<sup>e</sup> Includes elective day cases, non-elective short and long stays.
<sup>f</sup> includes software, additional server (if required), training, reporting costs and incentive costs (n=230 missing incentive cost) which was calculated if practices delivered four parts of the intervention (i.e. notes review, interprofessional discussion, clinical appointment with the patient and follow-up appointment unless clinically exempted).
<sup>g</sup> Includes GP, nurses, AHP, pharmacists, administrators across all modalities (face-to-face, virtual, home visits and administrative work).
<sup>h</sup> Includes outpatient appointments, admissions and AE visits.
<sup>i</sup> Sum of primary care, secondary care, intervention and prescribed medications costs

### 4.2 Outcomes

There was a small rate of missingness in primary care data at T_2_ (0.06%, n=1) and T_3_ (5.30%, n=100) but a greater rate of item missingness for self-reported SF12v2 responses (T_1_=6.82%, n=117; T_2_=30.50%, n=523; T_3_=33.76%, n=579). Due to intermittent missingness, total QALYs were available for 937 participants (54.64%) before imputation. Table 3 shows unadjusted mean utility and unadjusted VAS scores for both groups at each time point based on available data. During the trial period, participants in the intervention group accrued more QALYs (mean±SD) (0.653±0.170) than participants in the control group (0.615±0.167) resulting in an unadjusted incremental QALY of 0.024 (95% CI= 0.008 to 0.040, p=0.003). In general, mean utility scores at each time point were higher in the intervention group compared to the control group, though the unadjusted mean difference was only statistically significant at T_2_ (0.019, 95% CI= -0.002 to 0.036). Likewise for VAS scores, participants in the intervention reporting higher scores at each time point but a statistically significant mean difference was observed in T_2_ only (2.68, 95% CI= 0.163 to 5.200, p=0.037). In our analysis of the SF12v2 tool, we examined both the physical (PCS) and the mental (MCS) component scores, which range from 0 (lowest health) to 100 (highest health). We found a significant improvement in the MCS for the intervention arm at T_3_ by 1.244 (95% CI, 0.352 to 2.139). The mean±SD of the MCS in the intervention arm was 48.61±10.11 and was 47.22±10.64 in the control arm. No significant differences were observed between the two arms in the physical component scores.

### 4.3 Resource use and costs

Table 4 includes mean unadjusted costs for the intervention and control groups based on available cases. Mean primary care staff resource use cost (GPs, nurses, AHPs, pharmacists and administrators) was significantly higher in the intervention group (£729.77± £436.36) compared to the control group (£679.30 ± £427.93) with a mean difference of £50.47 (95%CI= £9.45 to £91.50). This difference appears to largely be the result of differences in pharmacists’ costs in each group (intervention=£66.03 vs. control=£22.22). Secondary care costs were also higher in the intervention group (£3949.22 ± £6488.84) compared to the control group (£3612.83 ± £5815.26), although there was no statistical evidence for a mean difference (95% CI= £-248.74 to £921.49, *p*=0.260). There was no significant difference in the cost of prescribed medications between the two groups (intervention=£1262.88 ± £1268.28 vs. control=£1323.40 ± £1436.72). Lastly, the overall NHS cost was higher in the intervention group compared to the control group (intervention= £5946.80 ± £5966.24, control = £5627.16 ± £6486.05) although there was no evidence of a statistically significant difference (95%CI= £-770.74 to £513.04). Mean costs for all cost categories for each HCP at each time point for both groups are shown in Supplementary Table S3 where we observed a slight reduction in administrative staff workload in the intervention group which seems to be offset by a slight increase in pharmacist administrative work at each time point. Social services resource use and costs are reported in Table 5. From a societal perspective, out of pocket data was available for only 56 participants in the control group (mean cost = £680.25 ± £1814.18) and for 92 participants in the intervention group (mean cost = £1599.89 ± £10779.28), due to low reporting of out-of-pocket expenses.

**Table 5:** Resource use and costs^a^ from a public perspective based on available cases.

| PSS and societal | N | IMPPP Intervention |  | N | Control |  | Incremental cost <sup>b</sup> (£)<br>(95% CI) | P value |
| --- | --- | --- | --- | --- | --- | --- | --- | --- |
|  |  | Mean<br>hours | Mean cost ±SD<br>(£) |  | Mean<br>hours | Mean cost ± SD<br>(£) |  |  |
| Care assistant | 393 | 1.08 | £24.76±454.68 | 327 | 3.22 | £74.12±855.49 | £-49.36 (-147.40 to 48.69) | 0.323 |
| Day care | 394 | 0.13 | £9.91±136.42 | 322 | 0 | £0 | £9.91 (-5.02 to 24.84) | 0.193 |
| Meals on Wheels | 393 | 0.23 | £1.26±16.39 | 326 | 1.04 | £5.69± 72.71 | £-4.43 (-11.85 to 2.99) | 0.241 |
| Home Help | 396 | 2.28 | £52.50±626.65 | 328 | 0.67 | £15.47±146.42 | £37.03 (-32.42 to 106.49) | 0.296 |
| Social worker | 392 | 0.59 | £29.66±463.15 | 324 | 0.18 | £8.97±103.53 | £20.69 (-31.50 to 72.89) | 0.437 |
| Out of pocket cost | 92 | - - - | £1599.89±10779.28 | 59 | - - - | £680.25±1814.18 | £919.64 (-1881.68 to 3720.95) | 0.518 |

<sup>a</sup> Costs and resource use from $T_1$ to $T_3$ . $S_2$ table lists unit costs <sup>b</sup> Incremental differences between intervention and control groups.

### 4.4 Economic analysis

While the intervention was associated with higher NHS costs, QALYs were nearly identical between groups. Compared to the control group, adjusted mean NHS costs were £511.17 significantly higher in the intervention group (95%CI= £73.22 to £949.13) whilst adjusted QALYs were just 0.006 higher (95%CI= -0.002 to 0.014) [Table 5]. Based on the iNMB result (£-419.37, 95%CI= £-920.37 to £81.91) and an ICER of £89,707/QALY, the intervention is not cost-effective at the current NICE WTP thresholds of £20,000 - £30,000 per QALY. Further, the CEAC suggests a 6.19% probability of the intervention being cost-effective at £20,000/QALY WTP (Figure 2 shows the probability of the intervention being cost-effective at different WTP thresholds). Compared to the control group, the intervention was more expensive and did not reduce the number of PIP indicators at T_3_ (Table 7) suggesting the usual care is dominant. From NHS perspective, it would cost £5225.25 per PIP indicator avoided. Figure 3 shows the CEAC which demonstrates that the intervention has a 10% probability of being cost-effective at different WTP thresholds per PIP indicator avoided.

**Figure 2:**
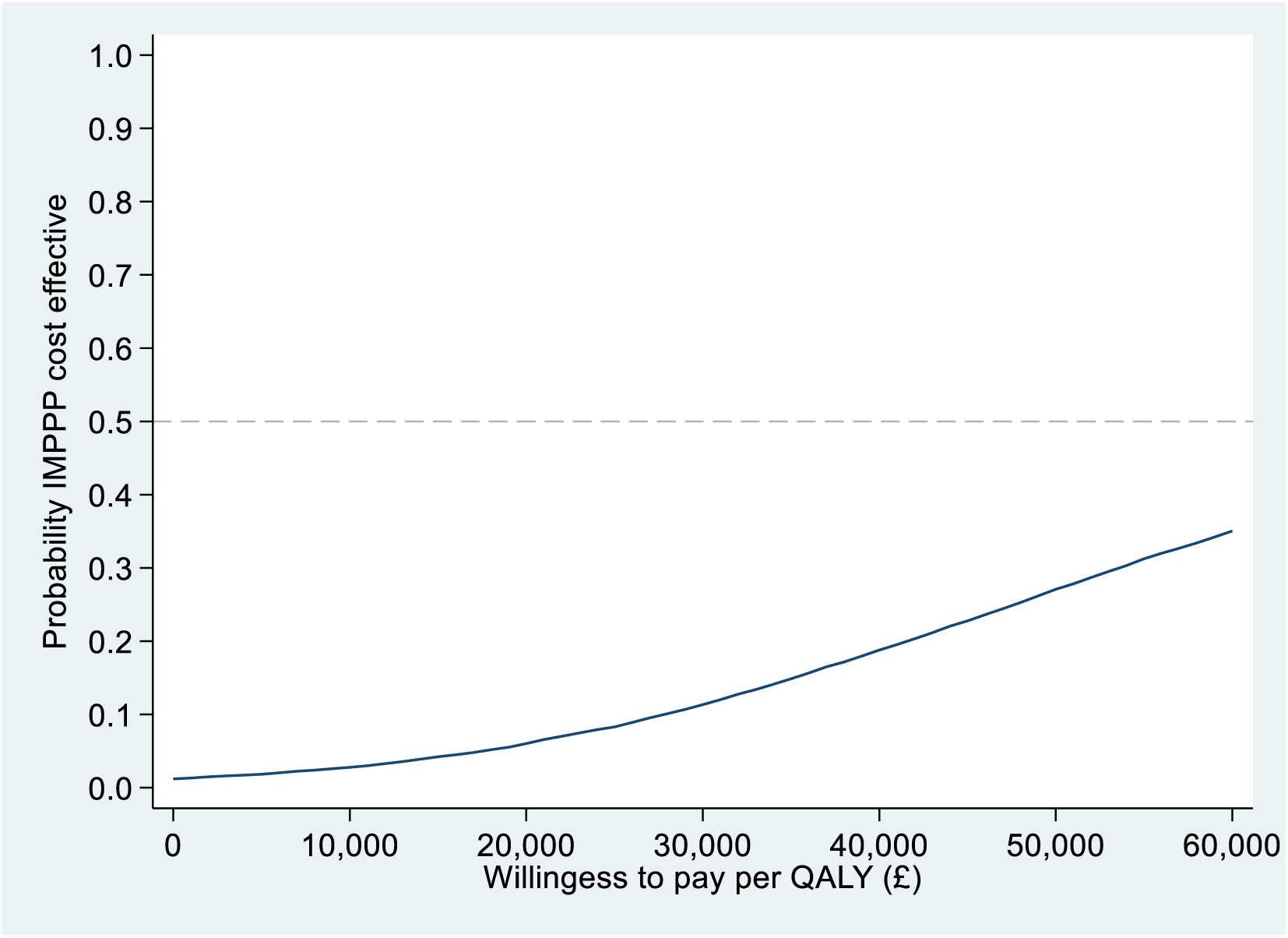
Cost Effectiveness Acceptability Curve (CEAC) from an NHS perspective.

**Figure 3:**
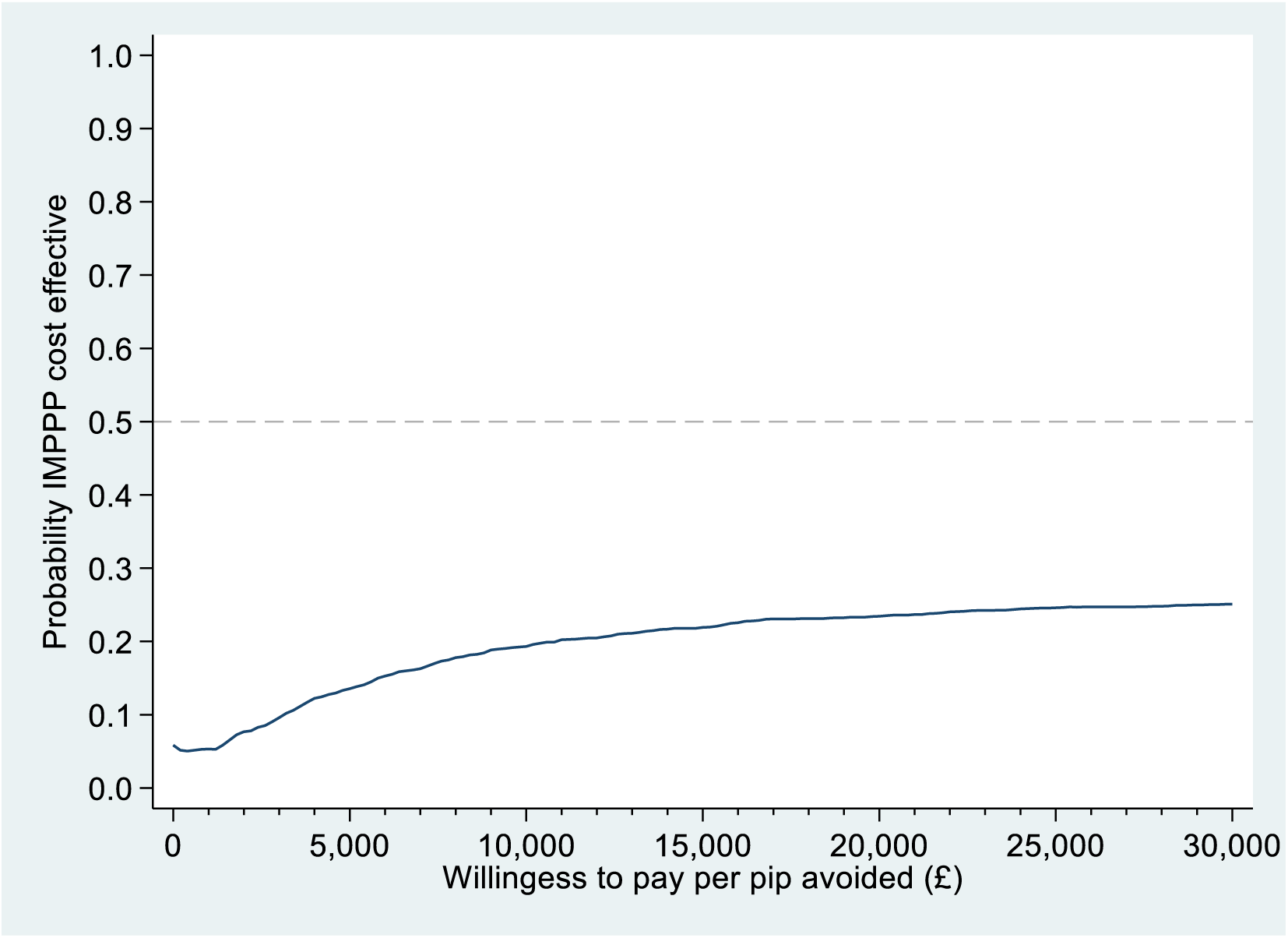
Probability of IMPPP cost per PIP indicator avoided.

### 4.5 Sensitivity Analysis

In the complete case analysis of costs and QALYs, NHS costs were lower in the intervention group by £138.07 though the difference was not statistically significant (95% CI= £-652.10 to £375.96). While participants in the intervention group accrued significantly more QALYs, the incremental difference was marginal (0.012, 95%CI= 0.004 to 0.021) (model 2, Table 6). Adjustments for IT systems and mental health issues yielded incremental cost differences between the intervention and usual care of £506.58 (95% CI= £68.55 to £944.62) and £508.43 (95% CI= £70.66 to £946.20), respectively, highlighting uncertainty in these estimates (models 3 and 4, Table 6). Similarly, the incremental QALY difference was small, at 0.004 (95% CI= -0.002 to 0.011) (models 3 and 4, Table 6).

**Table 6:**
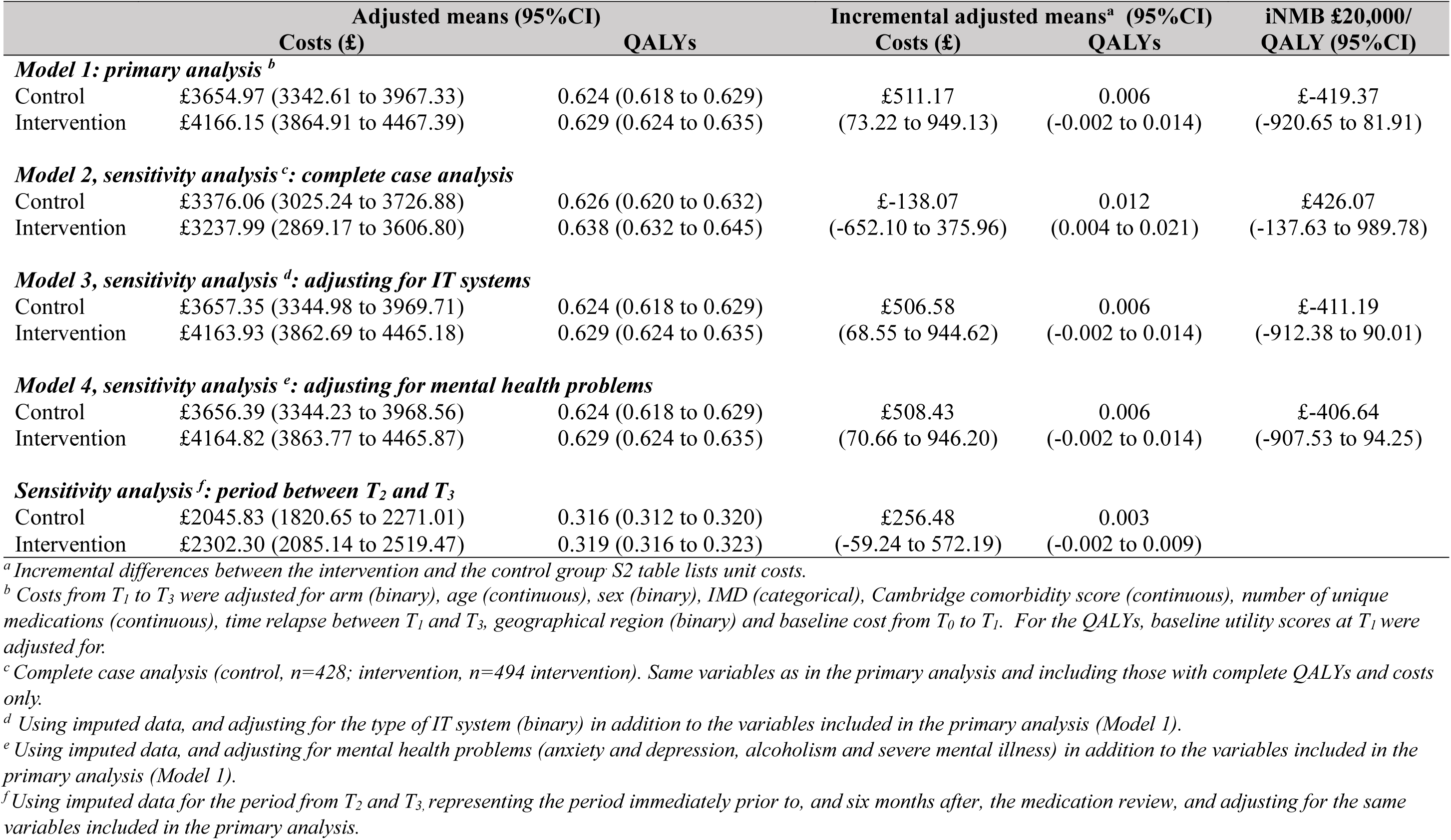
Primary, secondary and sensitivity analyses results.

**Table 7:**
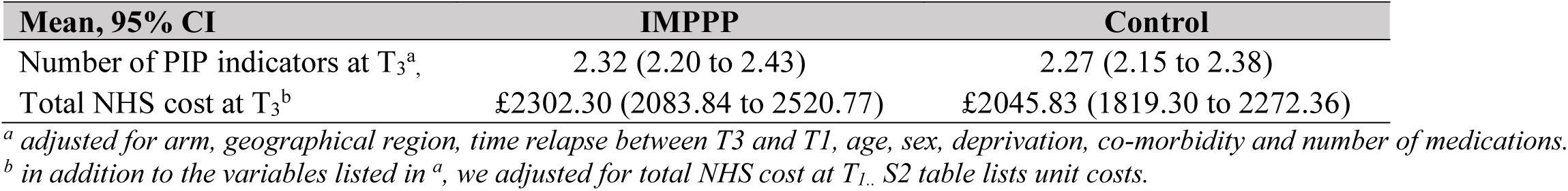
adjusted number of PIP indicators and total costs at T_3_.

When looking at the incremental cost between T_2_ and T_3_ – representing the period immediately prior to, and six months after the medication review, whilst adjusting for the same covariates as in the primary analysis, the intervention was more expensive although not significantly different than the control (£256.48, 95% CI = -£59.24 to £572.19). The model did not converge when estimating the net monetary benefit.

From a societal perspective, the mean ±SD total cost was £3201.65±£4257.59 (n=92) in the control arm and £4701.29 ±£11068.47 (n=134) in the intervention arm. The model did not converge until after excluding costs outliers (defined as above 95% percentile) in which the intervention was not cost-effective when compared to usual care with a mean increment cost difference of £175.53(95%CI= £-164.41 to £515.46).

## 5. Discussion

Results of our primary analysis show that there were no statistically significant differences in total mean costs or outcomes between the intervention and control patient groups from an UK NHS/ PSS perspective (Table 6). Results from the sensitivity analyses and societal perspective analysis are consistent with those of the primary analysis, with no statistically significant differences in mean costs or outcomes observed. Based on the results presented here, the IMPPP intervention is unlikely to be cost effective for commonly accepted ranges of willingness to pay per QALY of £20,000 to £30,000 (Figure 1). The heterogenous nature of the IMPPP study population may have reduced the likelihood of detecting a consistent clinical effect.

### 5.1 Strength and weaknesses

The trial had a number of strengths including pragmatic intervention design relevant to real world practice and policy, robust trial design, a larger sample size compared to similar trials (^14^) and the use of EHRs to improve the completeness of data. We also examined the completeness of two main outcome measures (EQ-5D-5L and SF12v2) in previous work in order to optimise detecting a difference in QoL at 6 months. Furthermore, the IMPPP trial shed light on a potential improvement in mental health following the intervention which can be attributed to a decrease in treatment burden although we are unable to conclude if this improvement is clinically significant (1, 20, 21). There are several limitations requiring consideration. Missing data in economic evaluation is quite common with a reported median proportion of trial participants with complete cost-effectiveness data of 63% (17). We used EHRs with a high rate of complete data on primary and secondary care service use (including prescribing), which mitigated the risk of recall bias associated with patient self-reported resource use tools (18). To address the high missingness in the self-reported SF-12v2, we carried out multiple imputation to avoid biased conclusions from available cases analysis (24). . We did conduct several sensitivity analyses, e.g. adjusting for mental health scores as measured by the Cambridge Multimorbidity Score, amongst others. Despite some evidence suggesting the sensitivity of SF-12v2 to prescribing practices on the physical component score of the SF12v2 (19), we could not replicate the same outcome in our study (20) and therefore cannot be certain about the sensitivity of the outcome. It is important to note that IMPPP patients represent a heterogeneous cohort with different clinical profiles which could explain our different results.

The time horizon of the economic evaluation was restricted to 6 months as we chose to mirror the primary study timeline, though we recognise that in many cases of economic evaluations of clinical studies authors construct a decision model to try and estimate costs and effects that occur beyond the period of data collection. The study follow-up period is insufficient to extrapolate robustly beyond the trial period using survival models as recommend by the ISPOR modelling good practice task force (26) and we believe that for our study a longer - term economic model would not be viable for three key reasons.

First, our study population included all patients seen in participating GP practices, meaning that our patient group had a wide range of clinical profiles. This included a wide variation in acute and chronic health care needs and age groups among other factors. The IMPPP study involved a single generic medication review that covered 15 clinical disease areas and included 118 PIP indicators. Previous studies of polypharmacy used more restrictive inclusion criteria and had more narrowly defined cohorts and outcome (e.g. medications errors) which may be more suitable to additional modelling. However, given the available data and resources we are unable to construct a model of longer-term impacts that could robustly represent the wide range of conditions and patient groups included in the IMPPP study, and so have restricted our analysis to the within study period. Second, given the 118 possible PIP indicator that were measured, there were a large number of possible medication changes that could be observed within the population and would also need to be modelled. While it may be possible to reduce the number of modelled changes to a set that is more manageable (for example, by condition) this would have required numerous assumptions about expected impacts of changes resulting from medication reviews. We believe the number of assumptions required would introduce too much uncertainty into the modelling process to make any conclusions valid. Furthermore, the intervention included further elements (training, performance feedback, etc) designed to improve broader aspects of pharmaceutical care, such as interprofessional engagement and patient-centredness, which have the potential to impact on relevant outcomes (e.g. quality of life, service use) but which could not be readily quantified and thus accounted for in a model. Finally, the trial was designed to incorporate a single medication review and, as discussed earlier, extending the time horizon to 12 months could have resulted in more than one review being undertaken in certain patient groups.

Due to logistical delays explained earlier, the time gap between T_1_ and T_2_ could have been associated with changes in the prescribing behaviour of physicians over time, although this was not evident in the sensitivity analysis. In addition, the introduction of a national policy on structured medications review (4–6) and the growth in recruiting clinical pharmacists in primary care may have influenced practices’ approach to medicines optimisation in the usual care arm, thus reducing any apparent effect. However, it is worth noting that recent research suggests sub-optimal delivery of these newer care models (21).

Because this was a GP-pharmacist-led intervention, pharmacists took on more of both the clinical and administrative workload, although this was offset to a degree by a reduction in administrative staff workload. Despite this increase in work, which is a necessary and integral part of the intervention, pharmacists and GPs perceived the intervention positively, including both the collaborative nature of the intervention and the enhanced training. (22).

### 5.2 Comparison with existing literature

The IMPPP trial stands out for recruiting a diverse cohort of patients that closely reflect real-world clinical settings. There are some methodological differences between the IMPPP trial and other relevant trials which may explain differing results of the effects of medication reviews targeting polypharmacy. Firstly, the perspective of analyses varied with some trials focusing on secondary care resource use only such as the OPERAM trial (23) and subsequently showing cost savings from a payer’s perspective.

Secondly, the IMPPP trial demonstrated a slight increase in administrative work amongst pharmacists and GPs and it remains unclear if other trials included this aspect in their costing e.g. the SPPiRE trial (24).-This administrative burden is a key factor that can significantly offset any savings from an intervention.

Thirdly, comparison of the IMPPP trial’s results with previous studies is challenging due to variations in sample sizes, participant characteristics and follow-up period. For instance, the SPPiRE trial (^24^) included 403 patients aged ≥65 years who were prescribed highly complex polypharmacy with ≥15 repeat medicines. The PINCER trial (25) (n=240) focused on a narrower range of PIP indicators as opposed to polypharmacy per se. The OPTICAL study (26) included 320 adults aged ≥65 years with ≥3 chronic conditions and ≥5 medications. The follow-up period between trials varied, from 6 months for IMPPP or SPPiRE (24) to nearer a year for the 3D trial (27), OPERAM (23), OPTICAL (26) and PINCER (25). Longer periods of follow-up may provide greater opportunity to detect longer-term outcomes relevant to medicines optimisation, including reduction in admissions, reduced prescribing, or improved quality of life.

Finally, it is also worth considering whether the outcomes assessed are sensitive to medication review interventions. For example, physical quality of life may be relatively hard to change through medication optimisation alone. Furthermore, there are no established means of applying costs to outcomes such as treatment burden, which may be more relevant to interventions that are not specific to a single disease or therapeutic area.

## 6. Conclusion

Health economic evaluation of the IMPPP trial showed that a complex intervention comprising medication review targeting polypharmacy, supported by informatics and other elements to enhance engagement, is not cost-effective. It highlights the difficulty of drawing firm conclusions when designing trials reflecting real-world settings, although also demonstrates the potential benefits of using routine data to following patients after medication review or other similar interventions. Future research might consider better approaches to maximise the economic time horizon despite limitations imposed by short clinical follow-up, and should develop improved quality of life instruments more sensitive to the effects of medication review unlike controlled RCTs with strict narrow inclusion criteria. Future research using observational data following patients after medication review is recommended.

## Statements and Declarations

The authors have declared no competing interests.

## Data availability

Requests for data sharing should be submitted to the Chief Investigator (RP) for consideration. Access to anonymised participant-level data might be granted following appropriate institutional review.

